# Human resources and healthcare infrastructure in Peru: A cross-sectional analysis from 2018 to 2024

**DOI:** 10.1101/2025.01.01.25319872

**Authors:** Luciana Bellido-Boza, David Villarreal-Zegarra, Paul Valdivia-Miranda

## Abstract

**Background:** The healthcare challenges from 2018 to 2024 in Peru focused on resource distribution, infrastructure gaps, and the impact of public policies on equity.

**Objective:** To describe the variations in the spatial distribution of healthcare professionals and available infrastructure in Peru between 2018 and 2024.

**Method:** Our design is observational. The Electronic Information Transfer System of Health Service Provider Institutions (IPRESS) and IPRESS Management Units (SETI-IPRESS) database was used between 2018 and 2024. Our study conducted a spatial visual analysis and a multiple linear regression mode

**Results:** Our study reveals an increase in IPRESS reporting data to SETI IPRESS in Peru, resulting in an increase in reported coverage from 2,255 institutions in 2018 to 3,560 in 2024. Additionally, significant growth has been identified in the average number of total clinics, beds, psychologists, medical technologists, obstetricians, pharmacists, and assistants available within IPRESS between 2018 and 2024. However, the average number of nurses and dentists decreased. Furthermore, a concentration of medical resources and infrastructure is observed in specific geographical areas, such as the departments of Lima and Lambayeque.

**Conclusions:** This study highlights the importance of health policies and strategies that could increase the number of care centers and improve the quality and equity in the distribution of health resources throughout Peru.

## BACKGROUND

In recent decades, equity in access to quality healthcare services has emerged as a central concern in public health policies globally, particularly in Latin America [1]. To reduce inequities in access, several Latin American countries have implemented policies that focus on expanding health coverage toward universal and equitable health care [2,3]. These policies have focused on reducing inequalities in the distribution of resources, the lack of trained personnel in remote areas, and the economic barriers still faced by the most vulnerable population [2].

A critical aspect of ensuring equity in a public healthcare system is the accessibility and geographical distribution of healthcare resources, including the availability of healthcare professionals and infrastructure [4]. The distribution of these resources tends to be inequitable, especially in low- and middle-income countries, because they have geographic areas with less access to services, fewer health professionals, and infrastructure with many limitations that characterize these so-called rural areas. Inequitable distribution can adversely affect the quality of clinical care and exacerbate health inequalities among the population [5].

Due to its fragmented nature, Peru’s health system faces many complexities, particularly in implementing public health strategies. This fragmentation has hindered coordination and efficiency in service delivery, especially in rural areas. Nevertheless, significant efforts have been made to improve access to quality health services, such as expanding the Comprehensive Health Insurance System (SIS, in Spanish) coverage and investing in health infrastructure in underserved areas [6,7].

This study aims to describe the spatial distribution variations of healthcare professionals and infrastructure in Peru from 2018-2024. Through this study, we aim to contribute to the understanding of healthcare resource distribution dynamics in Peru, providing a valuable tool for policymakers, health planners, and other stakeholders involved in developing practical and equitable health strategies at the national level.

## METHODS

### Design

The design was cross-sectional, and we used secondary data from the Electronic Information Transfer System database of Health Service Provider Institutions (SETI IPRESS). The National Superintendence of Health of Peru (SUSALUD) provided the data from January 2018 to January 2024.

### Setting

We conducted the study in Peru, a country characterized by its geographically diverse and dispersed population. Our analysis focused on human resources and healthcare infrastructure within Health Service Provider Institutions (IPRESS) nationwide from 2018 to 2024.

Peru needs a comprehensive system that integrates all health service data. In this context, our study utilized data collected from SETI IPRESS, one of the initial efforts to comprehensively gather information on the Peruvian health system, which is currently in the implementation phase. SETI IPRESS offers detailed information about the resources and infrastructure of IPRESS throughout Peru. The data collected from SETI IPRESS includes records on the number and distribution of healthcare professionals and available infrastructure within health facilities, such as medical equipment, hospital beds, and specialized care units.

Our analysis encompassed data from all available departments in Peru, allowing for a thorough and holistic assessment of health resource distribution. We examined public and private health establishments, considering various levels of care, from primary health services to specialized hospitals.

The SETI IPRESS data were systematically organized and analyzed to identify resource availability trends and assess potential healthcare access disparities over time. This approach provided an in-depth understanding of changes in Peru’s human resources and healthcare infrastructure during the study period.

The Peruvian health system has IPRESS at different levels of care, and each level must provide medical services according to the complexity and specialization required. Level I is for primary health care, minor medical centers, and centers focused on addressing issues that do not require high specialization. Level II includes hospitals and clinics of medium complexity, which can perform more complex diagnoses and offer emergency services and operating rooms. Level III care consists of high-complexity hospitals, such as national and specialized hospitals with advanced technology, intensive care units, and advanced diagnostic services.

### Participants

The participants in our study were the Health Service Provider Institutions (IPRESS) of the Peruvian healthcare system, both public and private. Information was collected on the number of healthcare professionals and available infrastructure for each month evaluated. The level of compliance with sending this information to SETI IPRESS varies according to the complexity level of each IPRESS. The differentiated compliance according to the level of complexity is presented in the attached figure, providing a clear view of adherence to documentation procedures in the various complexity categories of IPRESS (see Table 1). Because Level I has a limited SETI IPRESS implementation rate, only Levels II and III were considered.

**Table 1.** Distribution of compliance with data submission to SETI IPRESS by IPRESS in February 2024.

| Level of complexity | Total number of IPRESS | Number of IPRESS reporting to SETI IPRESS | % Compliance | % For level |
| --- | --- | --- | --- | --- |
| I-1 | 9349 | 1372 | 14.70% | 13% |
| I-2 | 6031 | 713 | 11.80% |  |
| I-3 | 4248 | 447 | 10.50% |  |
| I-4 | 455 | 119 | 26.20% |  |
| II-1 | 230 | 211 | 91.70% | 88% |
| II-2 | 84 | 80 | 95.20% |  |
| II-E | 249 | 204 | 81.90% |  |
| III-1 | 35 | 34 | 97.10% | 98% |
| III-2 | 14 | 14 | 100.00% |  |
| III-E | 10 | 10 | 100.00% |  |
| Uncategorized | 5052 | 22 | 0.40% |  |

### Variables

Our study utilized the variables in Table A of the SETI IPRESS, which reports the quantity of human resources in each IPRESS. Temporal information for each month per IPRESS was used, covering the period from January 2015 to January 2024.

#### Infrastructure outcomes

Our study will define the available infrastructure for healthcare services based on three variables: A) the total number of consulting rooms and the number of functional consulting rooms available within the IPRESS, B) the number of hospitalization beds available, and C) the number of operational and functioning ambulances available. Each of these variables was presented independently and self-reported by each IPRESS.

#### Healthcare human resource outcomes

We will define the outcome as the number of each available healthcare professional, specifically the total number of physicians, physicians during their social service (serums), physicians during the residency process, nurses, dentists, psychologists, nutritionists, medical technologists, obstetricians, pharmacists, assistants or auxiliary technicians, and other healthcare professionals. Self-reported information will be available as a sworn statement by the IPRESS for each type of healthcare professional.

### Procedure

Initially, the database coherence was reviewed using the quality control criteria provided by SUSALUD. Additionally, cases that did not correspond to reporting healthcare workers or infrastructure were eliminated. Subsequently, the data was exported to a STATA file for analysis.

### Analysis

First, we conducted a descriptive analysis grouped by year, which allowed us to identify the total number and average of healthcare workers and available infrastructure. We also included the total number of evaluated IPRESS, categorizing them by their level of complexity and the institution to which they belong. Second, we conducted a spatial visual representation by the department based on the average of healthcare workers and available infrastructure between the first and last measurements. Additionally, we presented data on the total number and average of healthcare workers and available infrastructure for each department, providing a clear view of the regional distribution of resources. Finally, we applied a multiple linear regression model to assess changes in the average of healthcare workers and infrastructure over time. For this analysis, a significance level of 0.05 and a confidence interval of 95% were established, ensuring the robustness and reliability of the results obtained.

### Ethical considerations

Our study does not involve ethical risks as it utilizes data from the SETI IPRESS, aggregated by month, and does not correspond to individual data where participants can be identified.

## RESULTS

### Description of the IPRESS

Our study identified that in January 2018, a total of 2,255 IPRESS reported their data in the SETI IPRESS, while for January 2024, this number increased to 3,560. Additionally, there was an increase in the total number of physicians between 2018 and 2024; however, the average number of physicians per IPRESS in 2018 was 19.1, while in 2024, it decreased to 15.3. Similarly, the average number of other healthcare professionals also reduced from 2018 to 2024, which could be explained by the increase in the number of primary healthcare facilities, which generally have lower complexity and, therefore, fewer staff.

Furthermore, although the number of available infrastructures increased between 2018 and 2024, the average number of health infrastructures per IPRESS decreased during this period. This could be due to the proportion of primary level IPRESS, which typically have less equipment, increasing during those years. The number of professionals and infrastructure available per year can be seen in Table 2.

**Table 2.**
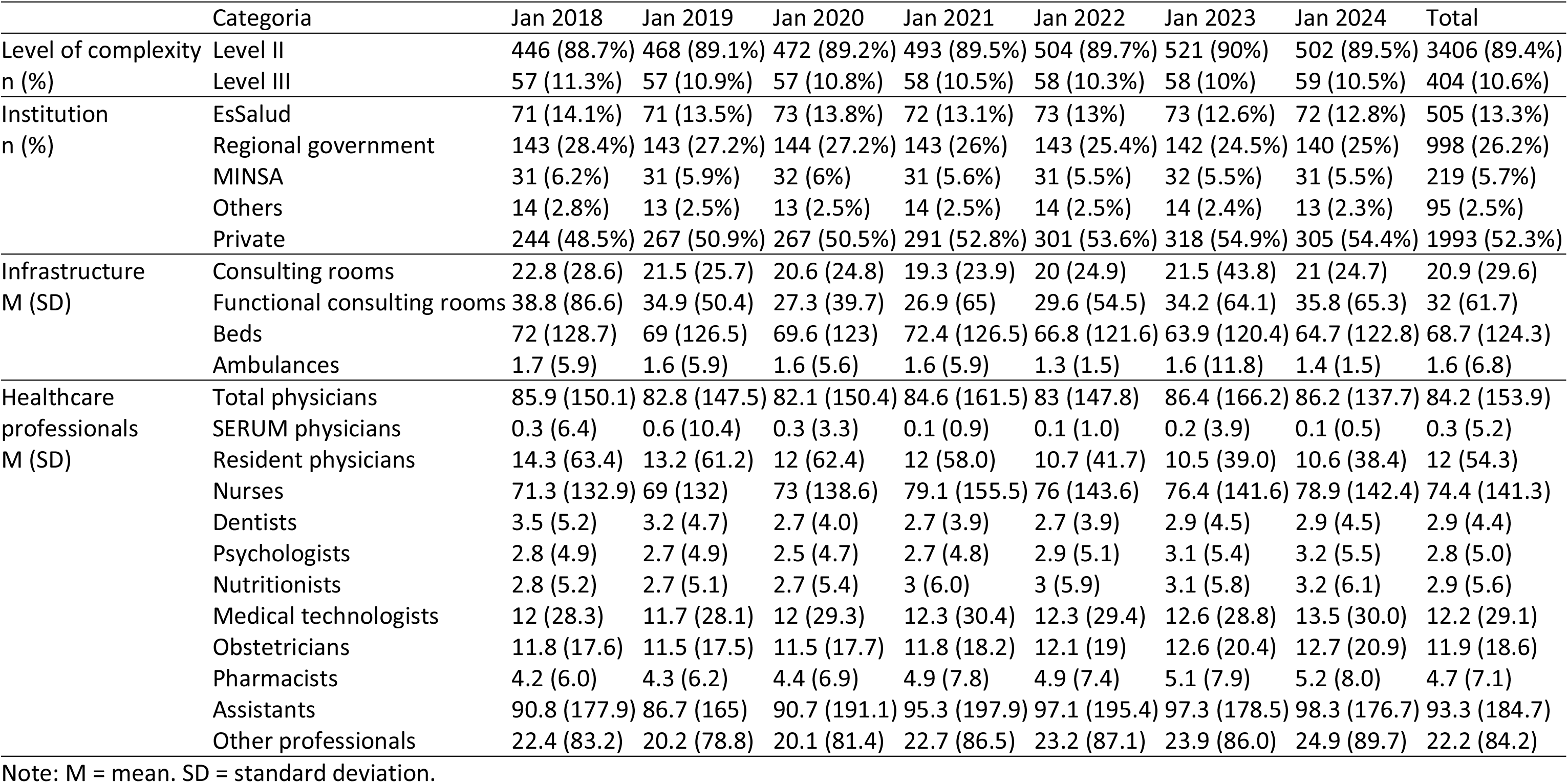
The average number of healthcare professionals and infrastructure available in Peru per IPRESS, reported to SETI IPRESS.

### Spatial Distribution of Healthcare Professionals and Infrastructure

Table 3 presents the spatial distribution of infrastructure and available healthcare professionals by department. Additionally, the comparison between 2018 and 2024 of the total number of physicians is illustrated in Figure 1, highlighting Lima and Lambayeque departments for having the highest average of physicians compared to other departments. This trend remained similar in the distribution of different types of healthcare professionals. Furthermore, the spatial distribution of the average number of beds also showed higher values in the Lima and Lambayeque departments than in the other departments, as seen in Figure 2. These data suggest a concentration of medical resources and infrastructure in some geographical regions, which could indicate inequalities in the distribution of healthcare services at the national level.

**Table 3.**
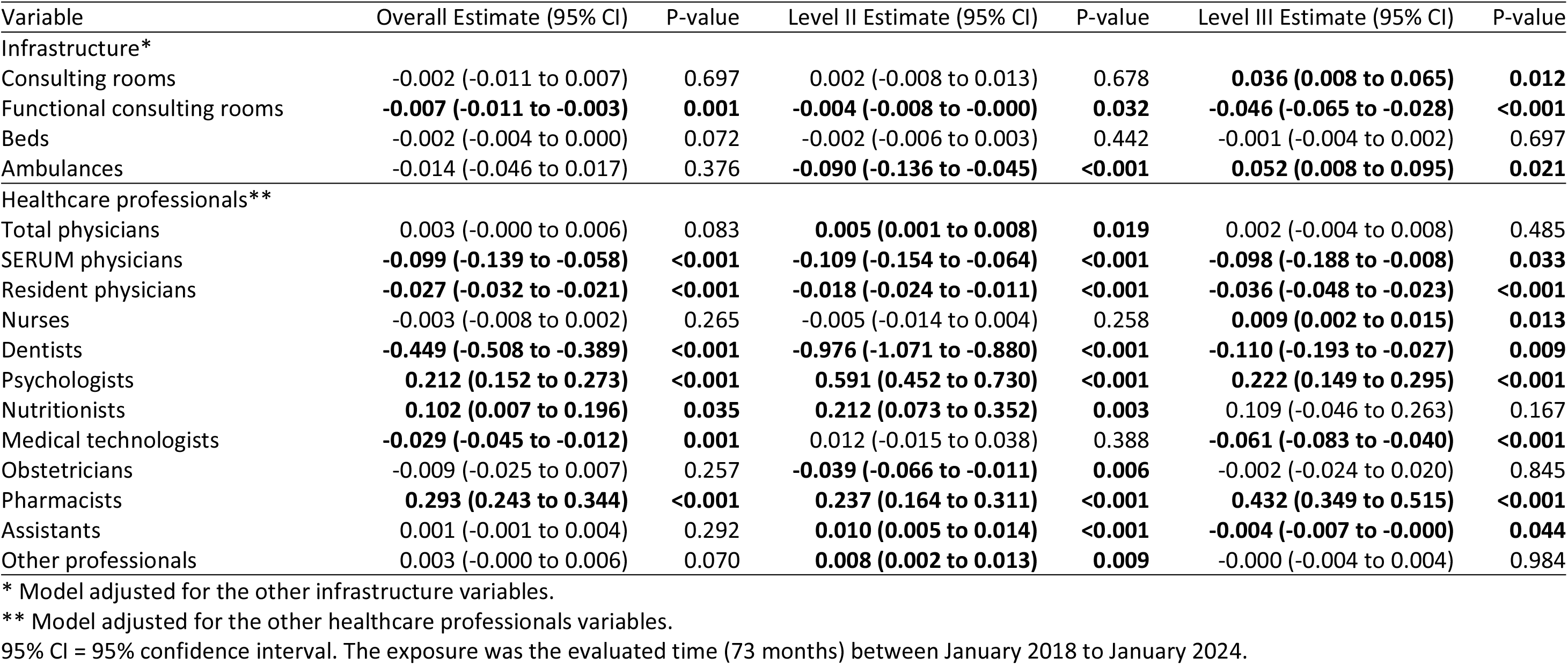
Linear regression models for available infrastructure and healthcare professionals by level.

| Variable | Overall Estimate (95% CI) | P-value | Level II Estimate (95% CI) | P-value | Level III Estimate (95% CI) | P-value |
| --- | --- | --- | --- | --- | --- | --- |
| Infrastructure* |  |  |  |  |  |  |
| Consulting rooms | -0.002 (-0.011 to 0.007) | 0.697 | 0.002 (-0.008 to 0.013) | 0.678 | **0.036 (0.008 to 0.065)** | **0.012** |
| Functional consulting rooms | **-0.007 (-0.011 to -0.003)** | **0.001** | **-0.004 (-0.008 to -0.000)** | **0.032** | **-0.046 (-0.065 to -0.028)** | **<0.001** |
| Beds | -0.002 (-0.004 to 0.000) | 0.072 | -0.002 (-0.006 to 0.003) | 0.442 | -0.001 (-0.004 to 0.002) | 0.697 |
| Ambulances | -0.014 (-0.046 to 0.017) | 0.376 | **-0.090 (-0.136 to -0.045)** | **<0.001** | **0.052 (0.008 to 0.095)** | **0.021** |
| Healthcare professionals** |  |  |  |  |  |  |
| Total physicians | 0.003 (-0.000 to 0.006) | 0.083 | **0.005 (0.001 to 0.008)** | **0.019** | 0.002 (-0.004 to 0.008) | 0.485 |
| SERUM physicians | **-0.099 (-0.139 to -0.058)** | **<0.001** | **-0.109 (-0.154 to -0.064)** | **<0.001** | **-0.098 (-0.188 to -0.008)** | **0.033** |
| Resident physicians | **-0.027 (-0.032 to -0.021)** | **<0.001** | **-0.018 (-0.024 to -0.011)** | **<0.001** | **-0.036 (-0.048 to -0.023)** | **<0.001** |
| Nurses | -0.003 (-0.008 to 0.002) | 0.265 | -0.005 (-0.014 to 0.004) | 0.258 | **0.009 (0.002 to 0.015)** | **0.013** |
| Dentists | **-0.449 (-0.508 to -0.389)** | **<0.001** | **-0.976 (-1.071 to -0.880)** | **<0.001** | **-0.110 (-0.193 to -0.027)** | **0.009** |
| Psychologists | **0.212 (0.152 to 0.273)** | **<0.001** | **0.591 (0.452 to 0.730)** | **<0.001** | **0.222 (0.149 to 0.295)** | **<0.001** |
| Nutritionists | **0.102 (0.007 to 0.196)** | **0.035** | **0.212 (0.073 to 0.352)** | **0.003** | 0.109 (-0.046 to 0.263) | 0.167 |
| Medical technologists | **-0.029 (-0.045 to -0.012)** | **0.001** | 0.012 (-0.015 to 0.038) | 0.388 | **-0.061 (-0.083 to -0.040)** | **<0.001** |
| Obstetricians | -0.009 (-0.025 to 0.007) | 0.257 | **-0.039 (-0.066 to -0.011)** | **0.006** | -0.002 (-0.024 to 0.020) | 0.845 |
| Pharmacists | **0.293 (0.243 to 0.344)** | **<0.001** | **0.237 (0.164 to 0.311)** | **<0.001** | **0.432 (0.349 to 0.515)** | **<0.001** |
| Assistants | 0.001 (-0.001 to 0.004) | 0.292 | **0.010 (0.005 to 0.014)** | **<0.001** | **-0.004 (-0.007 to -0.000)** | **0.044** |
| Other professionals | 0.003 (-0.000 to 0.006) | 0.070 | **0.008 (0.002 to 0.013)** | **0.009** | -0.000 (-0.004 to 0.004) | 0.984 |

\* Model adjusted for the other infrastructure variables.
\*\* Model adjusted for the other healthcare professionals variables.
95% CI = 95% confidence interval. The exposure was the evaluated time (73 months) between January 2018 to January 2024.

**Figure 1.**
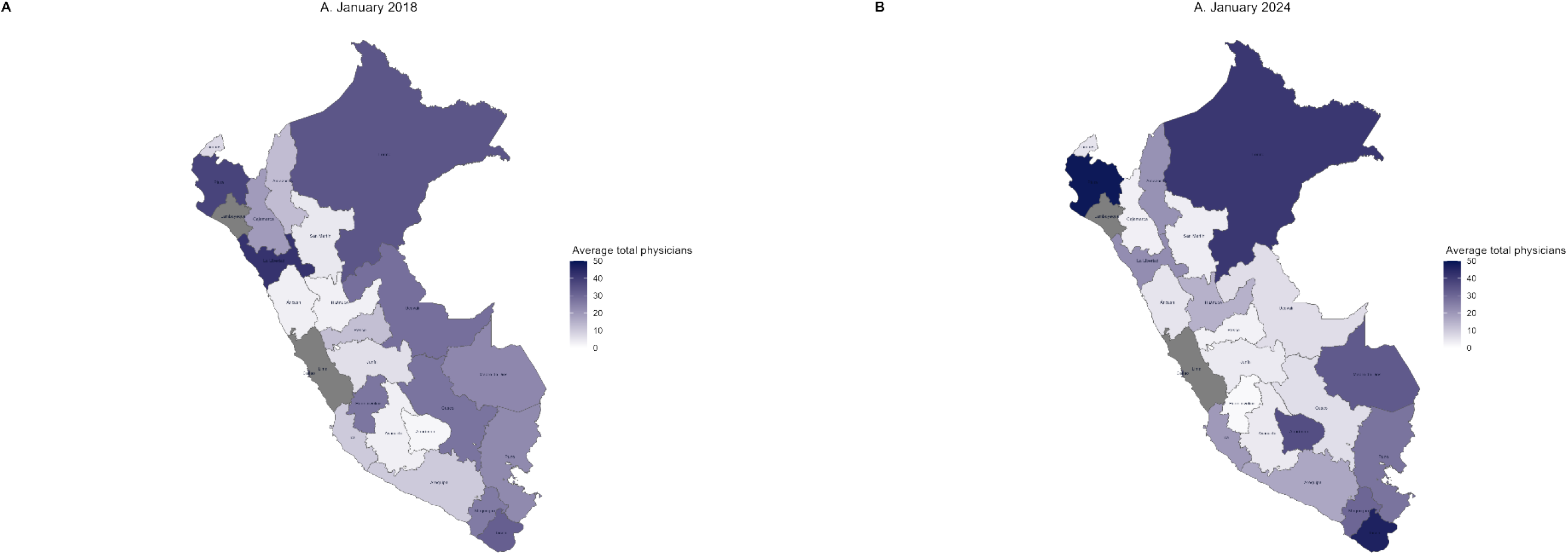
The average total physicians between January 2018 and January 2024, by department. Note: The departments shaded in gray correspond to those with more than 50 physicians on average.

**Figure 2.**
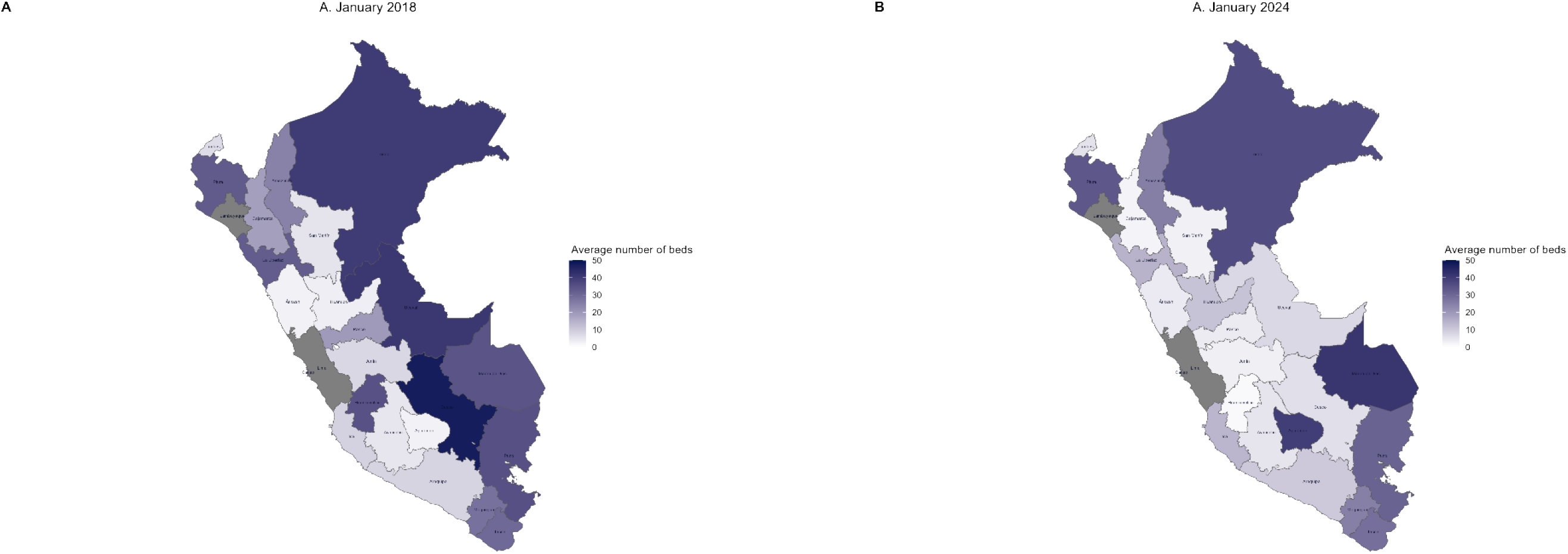
The average number of beds between January 2018 and January 2024, by department. Note: The departments shaded in gray correspond to those with more than 50 physicians on average.

### Changes in the average of healthcare professionals and infrastructure

Our study identified a significant monthly increase in the average number of total consulting rooms (β=0.006; p=0.001) and available beds (β=0.005; p=0.001) during the evaluated period. Additionally, we observed a significant monthly increase in the number of psychologists (β=0.01; p<0.001), medical technologists (β=0.011; p<0.001), obstetricians (β=0.008; p<0.001), pharmacists (β=0.028; p<0.001), and assistants (β=0.001; p=0.016) available within IPRESS during the same period. However, the average number of nurses (β=-0.004; p<0.001) and dentists (β=-0.031; p<0.001) decreased significantly each month (see Table 4).

## DISCUSSION

The observed increase in the number of IPRESS reporting data to SETI-IPRESS, from 2,255 in January 2018 to 3,560 in January 2024, reflects an improvement in the system’s coverage and capacity to capture information on healthcare services at the national level. However, this increase does not necessarily indicate a rise in operational health facilities but rather greater adherence to the SETI-IPRESS reporting requirements [8]. This distinction is important for correctly interpreting the reported expansion and evaluating the effectiveness of the policies implemented to improve transparency and accountability in the healthcare system.

Additionally, a decrease in the average number of healthcare professionals and infrastructure per IPRESS was identified. This phenomenon could be explained by the increase in the proportion of first-level IPRESS in 2024 compared to 2018. These first-level facilities generally require less specialized staff and infrastructure than higher-level institutions [1]. This highlights the need to allocate resources equitably to meet the growing nationwide healthcare service demand.

The analysis by level of complexity showed that IPRESS at levels II and III have a higher data reporting rate than those at level I. For instance, level II IPRESS reported 91.7% compliance, while level III reached 97.1%, indicating that these institutions have more resources and administrative staff available to meet reporting requirements [8]. This reveals that higher-level institutions have a greater capacity to manage data efficiently, which could reflect better infrastructure and access to trained professionals [4].

Finally, a significant increase in psychologists, pharmacists, and functional consulting rooms was observed between 2018 and 2024. These increases indicate an expansion in the availability of specialized services, which could improve access to and the quality of healthcare, particularly in mental health [9]. However, disparities persist in the distribution of these resources, with a greater concentration in urban areas such as Lima and Lambayeque.

Inequality in access to public health systems is a recurrent issue in Latin America, where individuals with higher economic status are more likely to access healthcare services [1,9,10]. In this context, public policies should focus on reducing these inequalities and promoting an adequate distribution of healthcare professionals and available infrastructure. Implementing robust information systems, such as SETI-IPRESS, can provide the necessary data to monitor equity in resource distribution and the effectiveness of health policies [8]. This would provide a powerful tool to identify potential disparities in the availability of healthcare professionals and infrastructure in Peru. Therefore, it is essential to emphasize the importance of solid health information systems that monitor and evaluate the equity and effectiveness of health interventions and policies.

Researchers invite healthcare intelligence teams and scholars to conduct analyses to identify inequalities in access to healthcare, using strategies such as concentration curves and Gini indices, which effectively measure inequalities in healthcare service utilization [4]. Such studies would help identify whether IPRESS located in low-income areas are typically underutilized compared to high-income areas, emphasizing the need for public policies to address these disparities.

### Strengths and limitations

One of the main strengths of the study is that it uses nationally reported data directly from IPRESS under sworn statements, minimizing the probability of information bias. However, the study presents some limitations. First, the implementation of SETI-IPRESS at the primary care level is low, as only 13% of IPRESS at this level reported data by February 2024. This limited implementation restricts the ability to generalize the findings to this level of care within the Peruvian healthcare system. Additionally, the lack of individual user information prevents assessing how patients move between different IPRESS, which is particularly relevant in a system centralized in Lima [4].

Moreover, geospatial analysis has proven effective in evaluating spatial accessibility and the distribution of healthcare personnel. This methodology accounts for the distribution of healthcare service providers and incorporates factors such as administrative boundary crossings and accessibility components related to infrastructure and transportation [4]. In Peru, there needs to be more information on the spatial distribution of health workers and available infrastructure. However, having such data would enable a better understanding of spatial variations in accessibility and could help effectively address inequalities in healthcare access, should they exist.

For future research, it would be beneficial to complement the data from SETI-IPRESS with external audits or field verifications to enhance the validity of the results. This would give policymakers a more reliable foundation for designing fairer and more equitable public health policies.

### Conclusions

Our study demonstrates a significant increase in IPRESS reporting data to the SETI-IPRESS system between 2018 and 2024, reflecting an improvement in the coverage of information on healthcare services at the national level. However, it is important to highlight that this increase does not necessarily indicate a rise in the creation of new healthcare facilities but rather greater adherence to the reporting requirements established by SETI-IPRESS. This improvement in system transparency is a crucial step toward greater accountability in providing healthcare services.

Additionally, important trends were identified in the availability of human resources and infrastructure at IPRESS. Despite the general increase in the number of consulting rooms, hospital beds, and professionals such as psychologists, obstetricians, and medical technologists, a decrease in the average number of nurses, dentists, and physicians per IPRESS was also observed. This reduction in healthcare personnel is particularly concerning, as it may affect the ability of facilities to provide comprehensive care, especially in rural and less developed areas of the country. These findings suggest that, while there are advances in the supply of certain specialized services, significant challenges remain in achieving equitable distribution of healthcare personnel.

The spatial analysis of resource distribution also reveals significant geographic inequalities, with a concentration of infrastructure and healthcare professionals in urban areas such as Lima and Lambayeque. This concentration may lead to an overload of healthcare services in these regions while other less populated and rural areas remain underserved. These disparities in resource distribution underscore the urgent need for more equitable health policies to ensure equal access to quality services across the entire Peruvian territory.

Finally, policymakers and health system planners need to consider implementing more robust information systems, such as SETI-IPRESS, to continuously monitor and evaluate the equity and effectiveness of health policies. Future research should focus on integrating external audits and geospatial analysis to identify healthcare access inequalities. This would provide valuable information for improving resource planning and ensuring a more equitable distribution of healthcare infrastructure in Peru [11].

## Data Availability

This research used freely accessible databases available on the SUSALUD open data platform. It should be noted that the data are available in an anonymized form and do not allow users to be identified. Therefore, the approval of an ethics committee will not be necessary, as we will not be working directly with the subjects included. The database was saved in Figshare (https://doi.org/10.6084/m9.figshare.26125492.v1).

## Acknowledgments

To the Dirección de Investigación of the Universidad Peruana de Ciencias Aplicadas for the support to complete this research work through the UPC-EXPOST-2025-1 incentive.

---

**Supplement 1.**
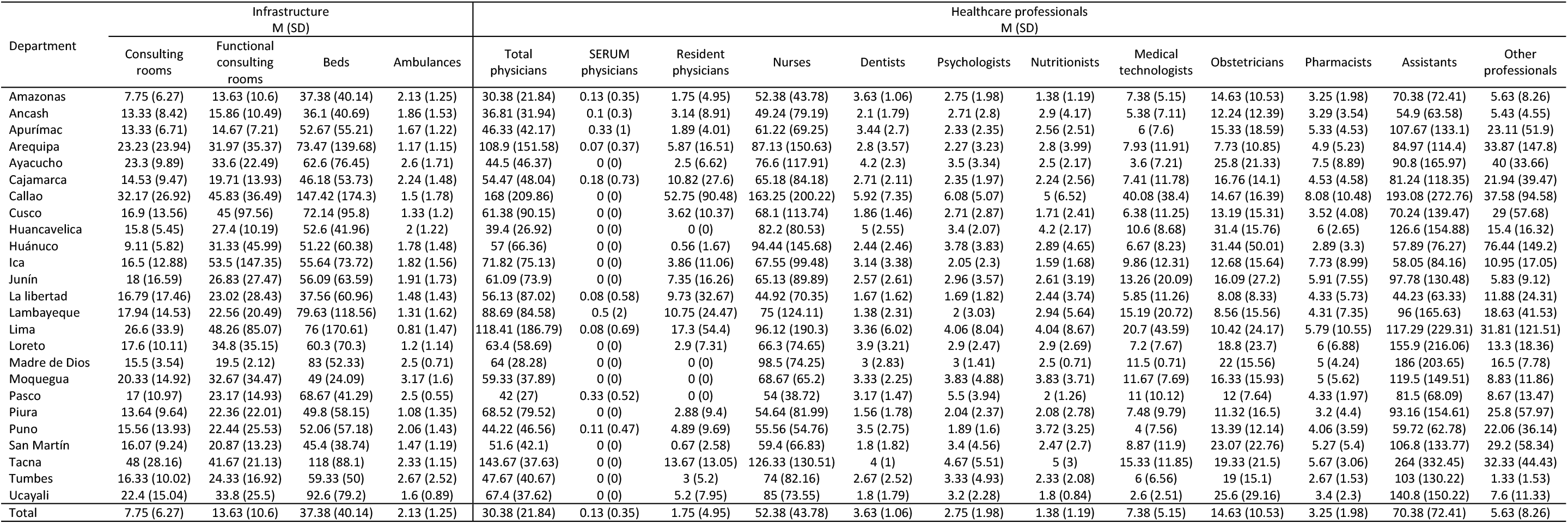
Averages and standard deviation of available infrastructure and healthcare professionals, by department, in January 2024.

## REFERENCES

[1] Frenz P, Titelman D. [Health equity in the world’s most unequal region: a challenge for public policy in Latin America]. Rev Peru Med Exp Salud Publica 2013;30:665–70.

[2] Dmytraczenko T, Almeida G. Toward universal health coverage and equity in Latin America and the Caribbean : evidence from selected countries. Washington: International Bank for Reconstruction and Development / The World Bank; 2015.

[3] Frenk J, Gómez-Dantés O. Health Systems in Latin America: The Search for Universal Health Coverage. Arch Med Res 2018;49:79–83. 10.1016/j.arcmed.2018.06.002.

[4] Love-Koh J, Griffin S, Kataika E, Revill P, Sibandze S, Walker S. Methods to promote equity in health resource allocation in low- and middle-income countries: an overview. Global Health 2020;16:6. 10.1186/s12992-019-0537-z.

[5] Cometto G, Boerma T, Campbell J, Dare L, Evans T. The Third Global Forum: framing the health workforce agenda for universal health coverage. Lancet Glob Health 2013;1:e324–325. 10.1016/S2214-109X(13)70082-2.

[6] Llanos Zavalaga LF, Castro Quiroz JA, Ortiz Fernández J, Ramírez Atencio CW, Llanos Zavalaga LF, Castro Quiroz JA, et al. Cuando crear sinergia no siempre es Salud: Análisis y propuesta en la evolución del Sistema de Salud en Perú. Revista Medica Herediana 2020;31:56–69. 10.20453/rmh.v31i1.3730.

[7] Sara DC, Carlos J. Lineamientos y estrategias para mejorar la calidad de la atención en los servicios de salud. Revista Peruana de Medicina Experimental y Salud Publica 2019;36:288–95. 10.17843/rpmesp.2019.362.4449.

[8] Superintendencia Nacional de Salud. Resolución de Superintendencia N.° 110-2023-SUSALUD/S. Perú: Superintendencia Nacional de Salud; 2023.

[9] Villarreal-Zegarra D, Al-kassab-Córdova A, Otazú-Alfaro S, Cabieses B. Socioeconomic and spatial distribution of depressive symptoms and access to treatment in Peru: A repeated nationwide cross-sectional study from 2014 to 2021. SSM - Population Health 2025;29:101724. 10.1016/j.ssmph.2024.101724.

[10] Al-Kassab-Córdova A, Silva-Perez C, Maguiña JL. Spatial distribution, determinants and trends of full vaccination coverage in children aged 12-59 months in Peru: A subanalysis of the Peruvian Demographic and Health Survey. BMJ Open 2022;12:e050211. 10.1136/bmjopen-2021-050211.

[11] Naylor KB, Tootoo J, Yakusheva O, Shipman SA, Bynum JPW, Davis MA. Geographic variation in spatial accessibility of U.S. healthcare providers. PLoS One 2019;14:e0215016. 10.1371/journal.pone.0215016.

